# Integrated Clinical and Network Pharmacology Study Reveals the Efficacy and Multi-Target Mechanism of Shenfu Injection in Septic Shock

**DOI:** 10.64898/2026.03.20.26348945

**Authors:** Bingyi Zhang, Yuan Tian, Qianqian Liu, Xiaoyan Zhou, Yahui Shi

## Abstract

**Background:** The high mortality of septic shock demands novel adjunctive therapies. Shenfu Injection (SFI), a traditional Chinese medicine, shows potential but its mechanism remains unclear.

**Methods:** We conducted an open-label, randomized trial in 80 patients with septic shock. Patients received standard care with or without adjunctive SFI for 7 days. The primary outcome was 28-day mortality. Key secondary outcomes included inflammatory markers, lactate clearance, and vasopressor duration. Concurrently, network pharmacology analyzed SFI’s bioactive components, predicted targets, and enriched pathways, with validation by molecular docking.

**Results:** The 28-day mortality was significantly lower in the SFI group (20.0% vs. 42.5%, P=0.030). SFI accelerated clinical improvement, evidenced by greater reductions in IL-6 and procalcitonin, higher 6-hour lactate clearance (35.2% vs. 18.5%, P<0.001), shorter vasopressor duration (48 vs. 72 hours, P<0.001), and more rapid SOFA score decline. Network pharmacology identified 145 SFI-septic shock common targets, with IL-6, SRC, and MAPK3 as central hubs. Pathway analysis revealed significant enrichment in TNF, PI3K-Akt, and IL-17 signaling pathways. Molecular docking confirmed strong binding of key SFI components (e.g., Ginsenoside Rh2) to core targets like IL-6.

**Conclusions:** Adjunctive Shenfu Injection reduces mortality and improves clinical recovery in septic shock, potentially through a multi-target mechanism involving modulation of inflammatory and cellular signaling pathways. This integrative study provides both clinical evidence and a mechanistic framework supporting SFI’s use. Clinical Trial Registration: Chinese Clinical Trial Registry, ChiCTR1800020435.

## Introduction

Septic shock, the most severe complication of sepsis, is characterized by profound circulatory, cellular, and metabolic disturbances, leading to a high risk of multiple organ dysfunction syndrome (MODS) and death [1,2]. Despite advances in antimicrobial therapy, early goal-directed resuscitation, and organ support, the morbidity and mortality rates of septic shock remain unacceptably high, posing a significant global health burden [3,4]. The pathophysiology involves a dysregulated host response to infection, culminating in a cytokine storm, endothelial damage, microcirculatory dysfunction, and cellular metabolic crisis [5,6]. This complexity makes single-target therapeutic strategies often insufficient, highlighting the need for novel approaches that can modulate the systemic inflammatory network.

In this context, traditional Chinese medicine (TCM), with its philosophy of holistic regulation and multi-target intervention, has shown complementary potential in the management of septic shock [7]. Shenfu Injection (SFI), a modern preparation derived from the classical TCM formula “Shenfu Tang” (composed of Red Ginseng and Aconite), is widely used in China for treating critical conditions characterized by “Yang depletion and collapse” [8,9]. It has been recommended by the Chinese guidelines for the emergency treatment of sepsis/septic shock for its potential benefits in stabilizing hemodynamics and improving outcomes [10]. Several clinical studies and meta-analyses have suggested that adjunctive therapy with SFI can elevate mean arterial pressure, reduce vasopressor dosage, improve microcirculation, and decrease short-term mortality in patients with septic shock [11,12].

However, the precise molecular mechanisms underlying these clinical benefits are not yet fully elucidated. SFI, like many other herbal injections, contains a multitude of bioactive compounds, making it challenging to deconvolute its pharmacological actions using conventional single-target research paradigms [13]. Network pharmacology, an approach that integrates systems biology and polypharmacology, provides a powerful framework for addressing this challenge. It allows for the prediction of potential active components, therapeutic targets, and key signaling pathways of multi-component medicines by constructing and analyzing biological networks [14]. This approach has been successfully applied to explore the mechanisms of various TCM formulations [15].

Therefore, while the clinical efficacy of SFI is increasingly recognized, a systematic exploration of its mechanism from a multi-component, multi-target network perspective is warranted. To bridge this gap, we conducted a study integrating a randomized clinical trial with a network pharmacology investigation. The clinical component aimed to evaluate the effects of SFI on inflammatory response, oxygen metabolism, and clinical outcomes in patients with septic shock. Concurrently, the network pharmacology component was designed to predict the active compounds, core targets, and pivotal pathways of SFI against septic shock, with the findings further validated by molecular docking. This combined strategy aims to provide both clinical evidence and a theoretical mechanistic basis for the application of SFI in septic shock management.

## Materials and Methods

### Clinical Study

#### Study Design and Ethics

This prospective, randomized, controlled, open-label trial was conducted in the Emergency Intensive Care Unit (EICU) of Hebei Cangzhou Integrated Traditional Chinese and Western Medicine Hospital between January 2023 and January 2025. The study protocol was reviewed and approved by the Ethics Committee of Hebei Cangzhou Integrated Traditional Chinese and Western Medicine Hospital (Approval No: CZX2023-KY-027.1). This trial was prospectively registered at the Chinese Clinical Trial Registry (Registration No: ChiCTR1800020435). Written informed consent was obtained from all patients or their legal surrogates prior to enrollment. The participant flow through the study is detailed in the Consolidated Standards of Reporting Trials (CONSORT) diagram (Figure 1). This study was conducted and reported in accordance with the CONSORT guidelines.

**Figure 1.**
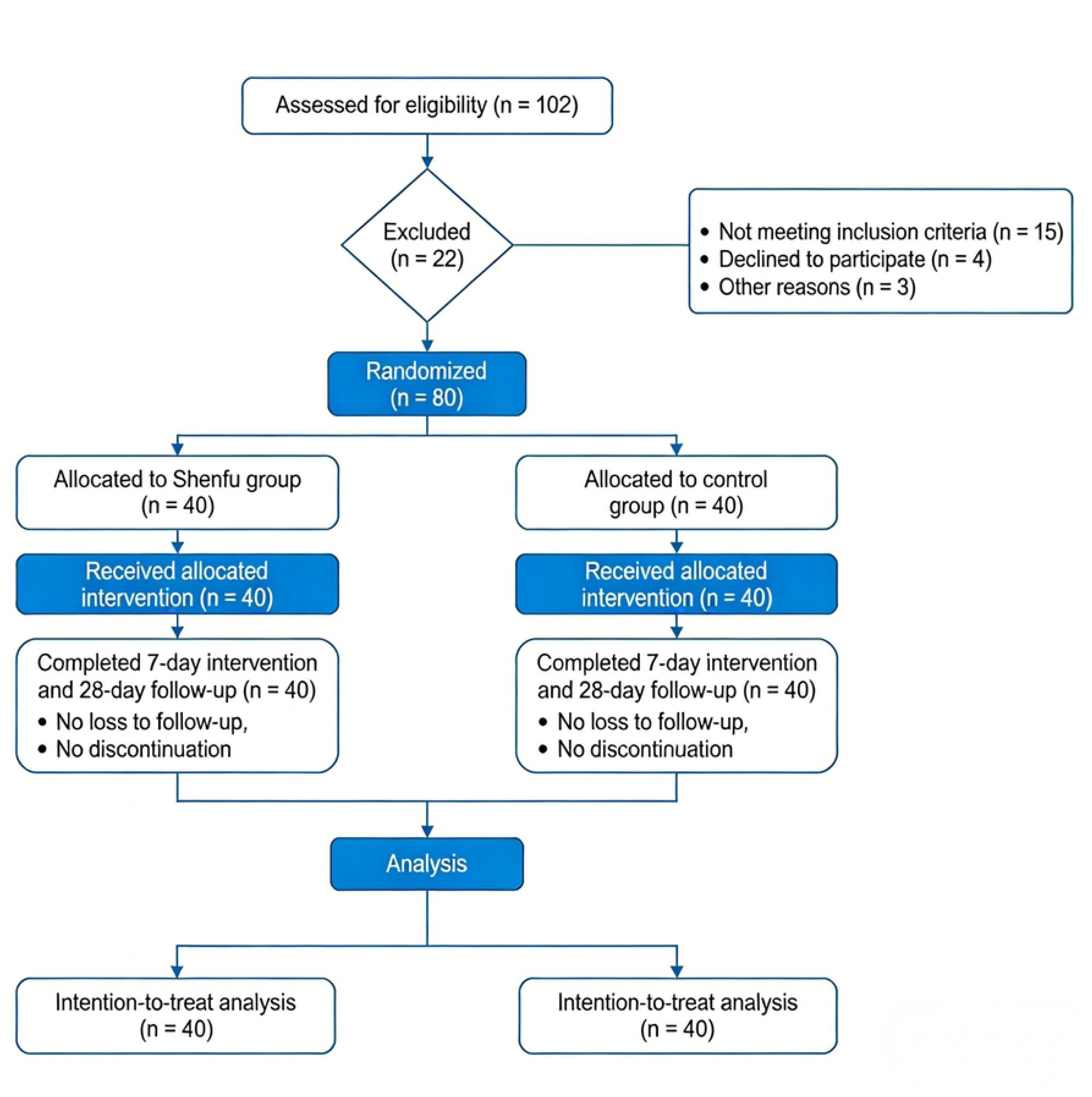
CONSORT flow diagram of the clinical trial.

#### Patients

Patients were eligible if they met the following criteria: (1) Diagnosed with septic shock according to the Sepsis-3 international consensus definitions: a confirmed or suspected infection, a Sequential Organ Failure Assessment (SOFA) score increase of ≥2 points, and the requirement for vasopressors to maintain a mean arterial pressure (MAP) ≥65 mmHg despite adequate fluid resuscitation, with a serum lactate level >2 mmol/L [16]; (2) Concurrently met the Traditional Chinese Medicine (TCM) diagnostic criteria for Yangqi Baotuo syndrome as defined by the “Guidelines for Clinical Research of Chinese Medicine in Treating Syncope and Collapse” (1989); (3) Aged between 18 and 65 years; (4) Provided written informed consent. Key exclusion criteria were: terminal malignancy, acute cardiovascular or cerebrovascular events, severe organ failure, coagulation disorders, known allergy to Shenfu Injection, pregnancy or lactation, and an expected survival time of less than 24 hours. Sample size calculation: Based on prior data indicating a 28-day mortality of approximately 45% in standard care, we estimated that Shenfu Injection could reduce mortality to 25%. With a two-sided alpha of 0.05 and 80% power (beta=0.2), the calculated sample size was 38 patients per group. Allowing for a 5% dropout rate, we enrolled 40 patients in each group.

#### Randomization and Interventions

Eligible patients were randomly assigned (1:1) to the Shenfu group or the control group using a computer-generated random sequence. Allocation was concealed with sequentially numbered, opaque, sealed envelopes. All patients received conventional standard therapy according to international guidelines, including early antimicrobial therapy, fluid resuscitation, vasopressor support (norepinephrine) to maintain MAP ≥65 mmHg, source control, and organ support as needed. Patients in the Shenfu group received an additional intravenous infusion of Shenfu Injection (National Medicine Permit Number: Z51020664; China Resources Sanjiu (Ya’an) Pharmaceutical Co., Ltd.; 60 ml diluted in 250 ml of 5% glucose solution) once daily for 7 consecutive days.

#### Outcome Measures

The primary outcome was all-cause mortality at 28 days. Secondary outcomes included: Inflammatory and metabolic markers: Serum levels of procalcitonin (PCT), C-reactive protein (CRP), interleukin-6 (IL-6), lactate, and lactate clearance rate at 6 hours; measured at baseline, 6 hours, and 7 days. Hemodynamic and oxygen parameters: PaO₂/FiO₂ ratio, central venous oxygen saturation (ScvO₂), and achievement of 6-hour early goal-directed therapy (EGDT) bundle targets. Clinical resource use: Duration of vasopressor use, mechanical ventilation, and ICU length of stay. Organ function: Changes in SOFA score and Acute Physiology and Chronic Health Evaluation II (APACHE II) score from baseline to day 3 and day 7. Safety: Incidence and type of adverse events, assessed for causality.

## Statistical Analysis

Analyses were performed using SPSS software (version 27.0) following the intention-to-treat principle. Continuous data are presented as mean ± standard deviation or median (interquartile range) and were compared using the independent samples t-test or Mann-Whitney U test, as appropriate. Categorical data are presented as numbers (percentages) and were compared using the Chi-square test or Fisher‘s exact test. Changes within groups over time were assessed with paired t-tests or Wilcoxon signed-rank tests. Factors associated with 28-day mortality were identified using univariate and multivariate logistic regression; variables with P < 0.10 in univariate analysis were entered into the multivariate model. A two-sided P value < 0.05 was considered statistically significant.

### Network Pharmacology and Molecular Docking Analysis

#### Screening of Active Components and Targets

Active components of Shenfu Injection (from Panax ginseng and Aconitum carmichaelii) were retrieved from the TCMSP database (http://tcmspw.com/tcmsp.php), filtered by oral bioavailability (OB) ≥ 30% and drug-likeness (DL) ≥ 0.18. Their canonical SMILES structures from PubChem were submitted to SwissTargetPrediction (http://www.swisstargetprediction.ch/) to predict putative targets (Homo sapiens, probability > 0). Targets were standardized to official gene symbols via UniProt (https://www.uniprot.org/).

#### Identification of Disease Targets and Common Targets

Septic shock-related targets were retrieved from the GeneCards database (https://www.genecards.org/) using the keyword “septic shock”. To ensure the relevance of targets, only genes with a relevance score greater than the median were retained for subsequent analysis. The intersection between drug component targets and disease targets was identified as potential therapeutic targets.

#### Protein-Protein Interaction (PPI) Network Construction and Analysis

Common targets were imported into the STRING database (https://string-db.org/, version 11.5) to construct a PPI network (species: Homo sapiens, minimum required interaction score: medium confidence > 0.400). The network file was analyzed and visualized in Cytoscape (version 3.9.1). Hub targets were identified based on high betweenness centrality.

#### GO and KEGG Pathway Enrichment Analysis

Gene Ontology (GO) and Kyoto Encyclopedia of Genes and Genomes (KEGG) pathway enrichment analyses for the common targets were performed using the DAVID database (https://david.ncifcrf.gov/, version 6.8). Terms with a Benjamini-Hochberg adjusted P value < 0.05 were considered significantly enriched.

#### Construction of the “Drug-Component-Target-Pathway” Network

An integrative network linking herbs, active components, targets, and enriched pathways was constructed and visualized using Cytoscape to elucidate the multi-scale intervention profile.

#### Molecular Docking Validation

The three-dimensional structures of core target proteins were obtained from the RCSB Protein Data Bank (PDB). Structures of key active components were prepared from TCMSP. Molecular docking simulations were performed using AutoDock Vina after preparing receptor and ligand files with AutoDockTools. Binding affinity was evaluated by the calculated binding energy (kcal/mol).

## Results

### Clinical Study Results

#### Baseline Characteristics of Patients

A total of 80 patients diagnosed with septic shock were enrolled and randomly assigned to either the Shenfu group (n=40) or the control group (n=40). The baseline demographic and clinical characteristics of the two groups are summarized in Table 1. There were no statistically significant differences between the two groups in terms of age, sex, body mass index, comorbidity burden (Charlson Comorbidity Index), source of infection, pathogen type, disease severity scores (APACHE II and SOFA), key physiological and laboratory values at enrollment (including mean arterial pressure, heart rate, serum lactate, procalcitonin, and C-reactive protein), or the time from ICU admission to intervention (all P > 0.05). This indicates that the two groups were well-balanced at baseline.

**Table 1.**
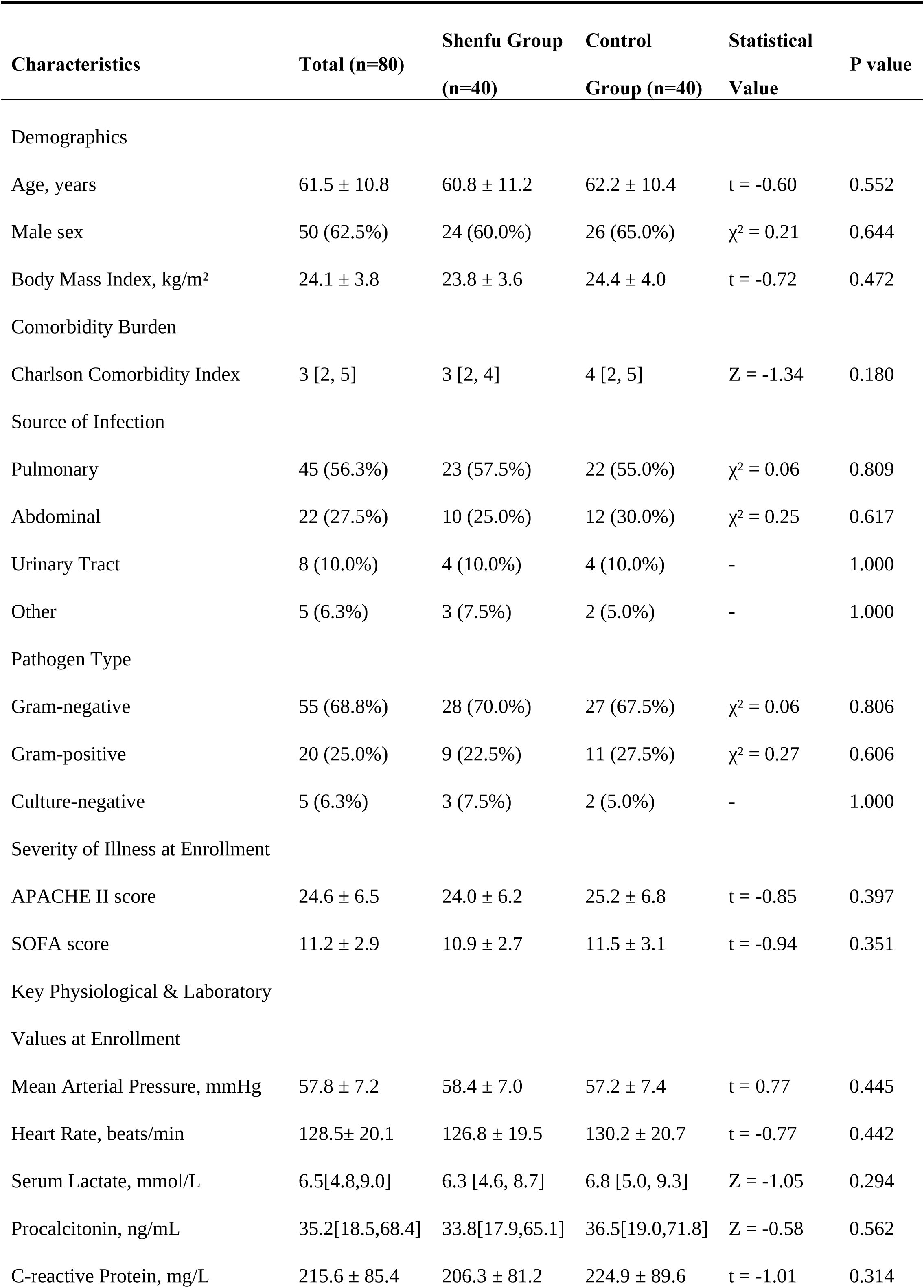
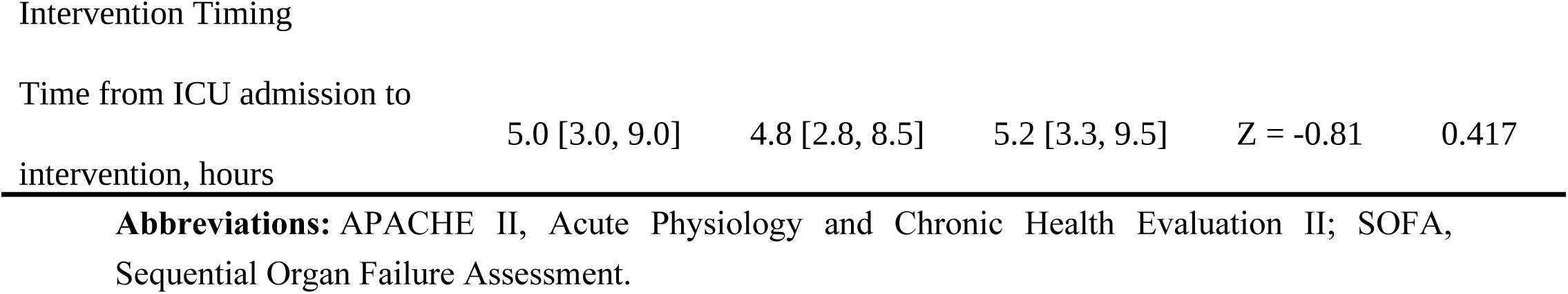
Baseline Characteristics and Comparability of Patients with Septic Shock.

#### Inflammatory and Oxygen Metabolism Indicators

The dynamic changes in inflammatory and oxygen metabolism indicators before and after treatment are shown in Table 2. Inflammatory Indicators: At 6 hours after treatment, the Shenfu group showed a more pronounced reduction in PCT and IL-6 levels compared to the control group (Intergroup P = 0.045 and 0.038, respectively). By day 7, levels of PCT, CRP, and IL-6 in the Shenfu group were significantly lower than those in the control group (all Intergroup P < 0.001). Oxygen Metabolism Indicators: At 6 hours, the reduction in serum lactate was greater in the Shenfu group than in the control group (Intergroup P = 0.002), and the 6-hour lactate clearance rate was significantly higher in the Shenfu group (35.2% ± 12.8% vs. 18.5% ± 15.4%, P < 0.001). The central venous oxygen saturation (ScvO₂) at 6 hours was also higher in the Shenfu group (Intergroup P = 0.003). At day 7, both serum lactate and ScvO₂ in the Shenfu group showed superior improvement compared to the control group (both Intergroup P < 0.01). The PaO₂/FiO₂ ratio was significantly higher in the Shenfu group at day 7 (Intergroup P < 0.001).

**Table 2.**
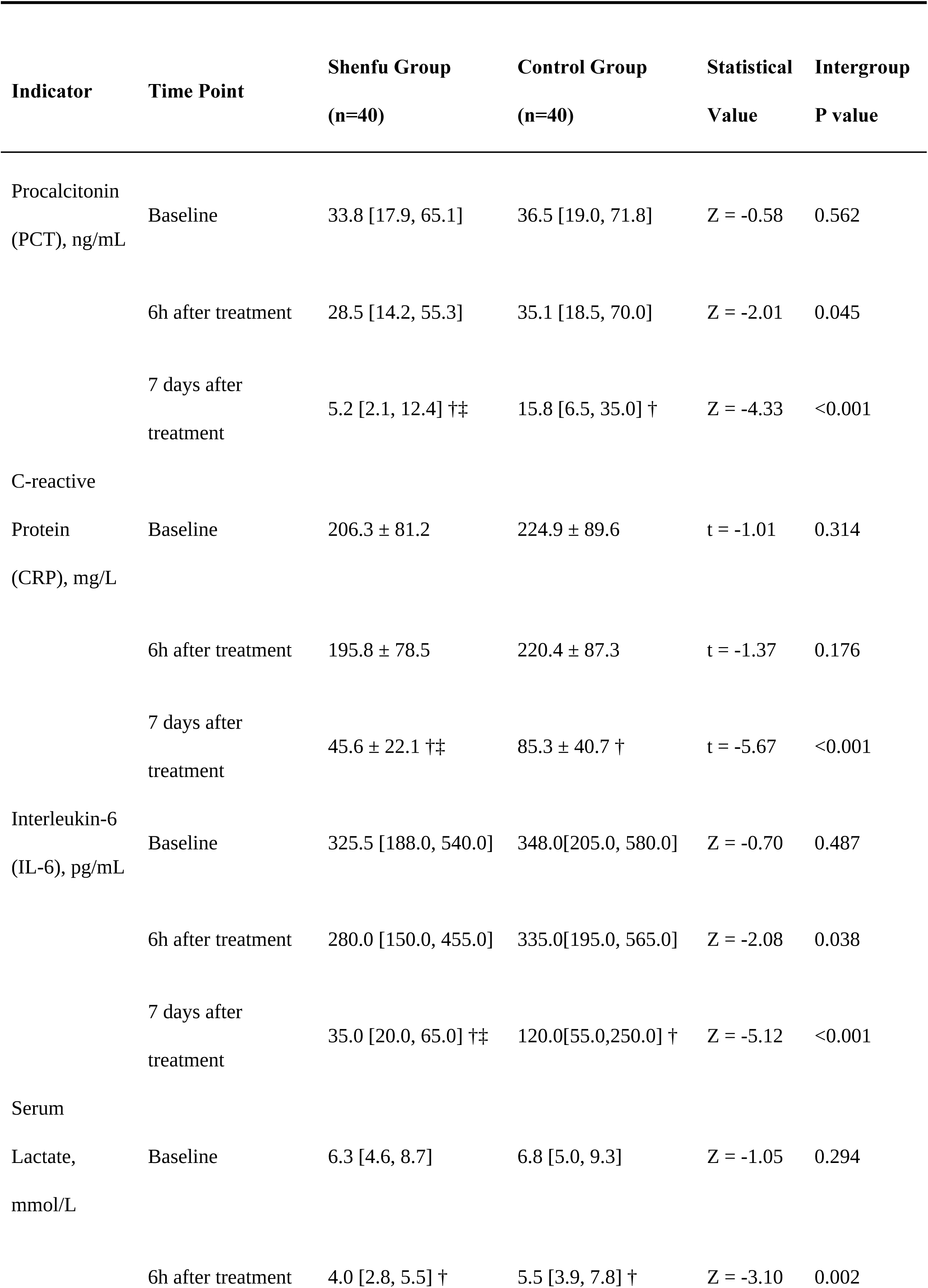
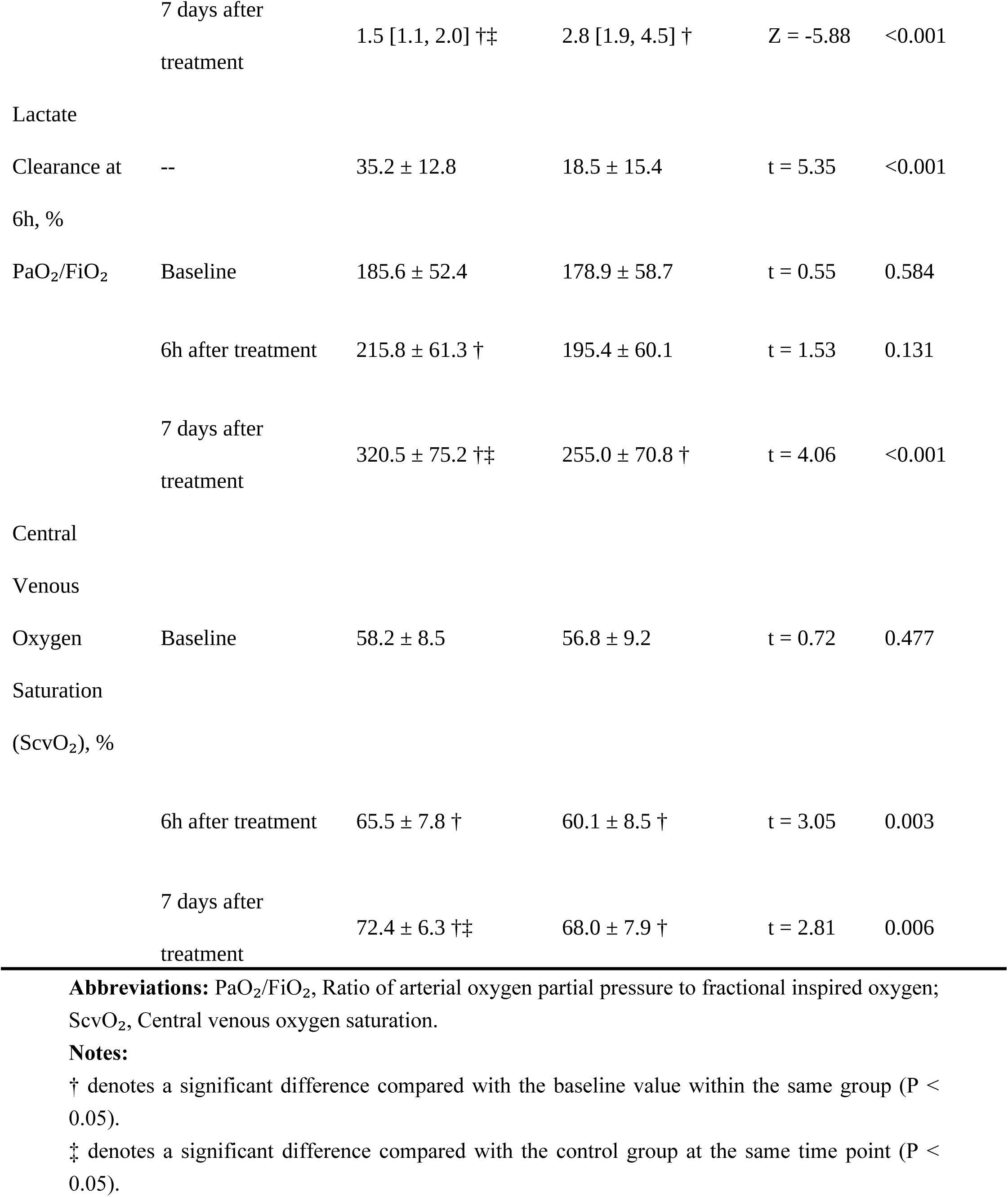
Comparison of Inflammatory and Oxygen Metabolism Indicators Before and After Treatment.

#### Clinical Outcomes and Safety

The primary and secondary clinical outcomes are presented in Table 3. Primary Endpoint: The 28-day all-cause mortality was 20.0% (8/40) in the Shenfu group, which was significantly lower than the 42.5% (17/40) in the control group (P = 0.030). Secondary Endpoints: Compared to the control group, the Shenfu group had a shorter median duration of vasopressor use (48 vs. 72 hours, P < 0.001), a shorter median duration of mechanical ventilation (96 vs. 144 hours, P = 0.002), and a shorter median length of ICU stay (7.0 vs. 10.0 days, P = 0.003). The improvement in organ function, as assessed by the ΔSOFA score from baseline to day 3 and day 7, was significantly greater in the Shenfu group (both P < 0.001). The ΔAPACHE II score from baseline to day 7 also showed a greater reduction in the Shenfu group (P < 0.001). A higher proportion of patients in the Shenfu group achieved the 6-hour EGDT bundle targets (80.0% vs. 55.0%, P = 0.017), and the time to achieve a mean arterial pressure ≥65 mmHg was shorter (120 vs. 180 minutes, P = 0.002). Safety: The overall incidence of adverse events did not differ significantly between the Shenfu group (15.0%) and the control group (12.5%) (P = 0.745). No serious adverse events related to the study treatment were reported.

**Table 3.** Comparison of Clinical Outcomes and Secondary Endpoints.

| Outcome Measures | Shenfu Group<br>(n=40) | Control Group<br>(n=40) | Statistical<br>Value | P value |
| --- | --- | --- | --- | --- |
| Primary Clinical Endpoint |  |  |  |  |
| 28-day all-cause mortality, n (%) | 8 (20.0%) | 17 (42.5%) | $\chi^2 = 4.71$ | 0.030 |
| Secondary Endpoints |  |  |  |  |
| Duration of vasopressor use, hours | 48 [24, 72] | 72 [48, 120] | $Z = -3.82$ | <0.001 |
| Mechanical ventilation duration, hours | 96 [48, 144] | 144 [96, 192] | $Z = -3.12$ | 0.002 |
| Length of ICU stay, days | 7.0 [5.0, 10.0] | 10.0 [7.0, 14.0] | Z = -2.95 | 0.003 |
| Organ Function Scores |  |  |  |  |
| ΔSOFA score (Baseline to Day 3) | -3 [-4, -2] † | -1 [-2, 0] | Z = -5.10 | <0.001 |
| ΔSOFA score (Baseline to Day 7) | -6 [-7, -4] † | -3 [-4, -1] † | Z = -5.88 | <0.001 |
| ΔAPACHE II score (Baseline to Day 7) | -10.5 ± 4.2 † | -5.8 ± 3.9 † | t = 5.25 | <0.001 |
| Early Goal-Directed Therapy |  |  |  |  |
| Metrics |  |  |  |  |
| Achievement of 6-h EGDT bundle, n (%) | 32 (80.0%) | 22 (55.0%) | $\chi^2 = 5.70$ | 0.017 |
| Time to achieve MAP ≥65 mmHg (min) | 120 [90, 180] | 180 [120, 240] | Z = -3.05 | 0.002 |
| Safety Outcomes |  |  |  |  |
| Total adverse events, n (%) | 6 (15.0%) | 5 (12.5%) | $\chi^2 = 0.11$ | 0.745 |
| • Cardiac arrhythmia | 2 (5.0%) | 1 (2.5%) | - | 1.000 |
| • Elevated liver enzymes | 3 (7.5%) | 2 (5.0%) | - | 1.000 |
| • Allergic reaction | 1 (2.5%) | 2 (5.0%) | - | 1.000 |

**Abbreviations:** Δ, change from baseline; SOFA, Sequential Organ Failure Assessment; APACHE II, Acute Physiology and Chronic Health Evaluation II; EGDT, Early Goal-Directed Therapy; MAP, Mean Arterial Pressure; ICU, Intensive Care Unit.
**Notes:**
† denotes a significant change from baseline within the group (P < 0.05).

#### Analysis of Factors Associated with 28-day Mortality

Univariate and multivariate logistic regression analyses were performed to identify factors associated with 28-day mortality (Table 4). In univariate analysis, higher APACHE II score, higher SOFA score, higher serum lactate at enrollment, a Charlson Comorbidity Index ≥3, and treatment without Shenfu injection were significantly associated with increased odds of mortality. In the multivariate model, after adjustment for other factors, APACHE II score (adjusted OR = 1.10, 95% CI: 1.01-1.20, P=0.024) and serum lactate at enrollment (adjusted OR = 1.22, 95% CI: 1.04-1.44, P=0.016) remained independent risk factors for mortality. Treatment with Shenfu injection showed a strong trend towards being an independent protective factor, though it did not reach the conventional threshold for statistical significance in this model (adjusted OR = 0.39, 95% CI: 0.14-1.11, P=0.078).

**Table 4.**
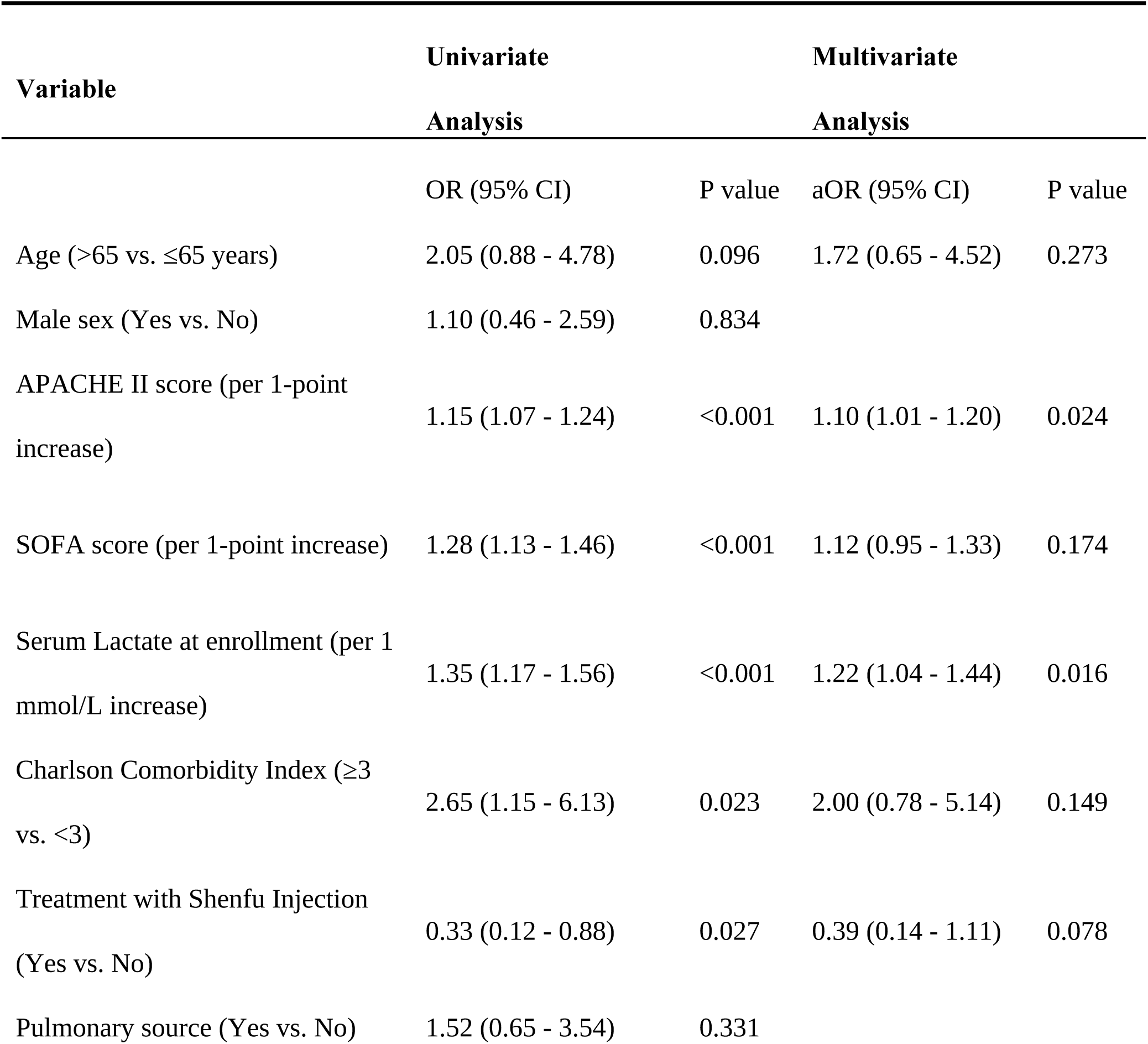
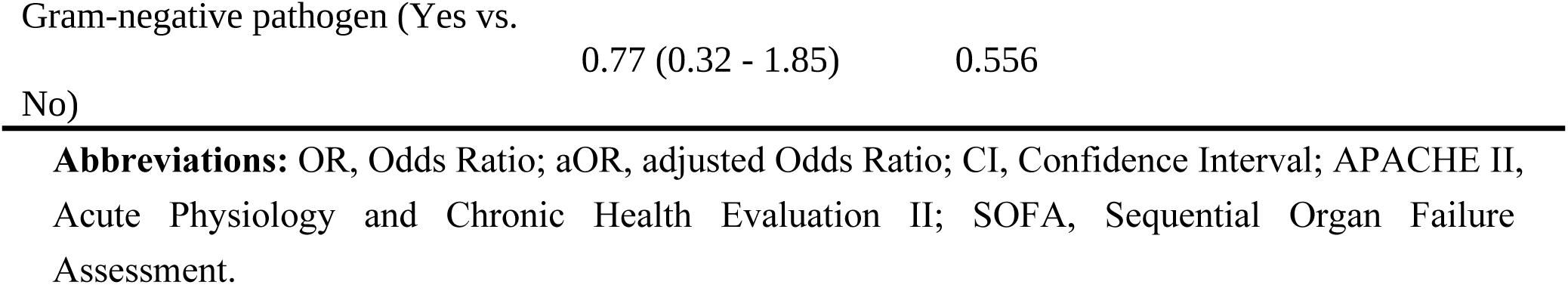
Univariate and Multivariate Logistic Regression Analyses of Factors Associated with 28-day Mortality in Patients with Septic Shock.

## Network Pharmacology Analysis Results

### Active Components and Potential Targets Screening

A total of 25 active components in Shenfu Injection (4 from Red Ginseng and 21 from Aconite) were identified from the TCMSP database after screening with the criteria of oral bioavailability (OB) ≥ 30% and drug-likeness (DL) ≥ 0.18 (Table 5). Through the SwissTargetPrediction database, 472 potential targets for these components were obtained. Simultaneously, 2,113 targets related to septic shock were retrieved from the GeneCards database. The intersection of these two target sets yielded 145 common targets, which were identified as the potential therapeutic targets of Shenfu Injection for septic shock.

**Table 5.**
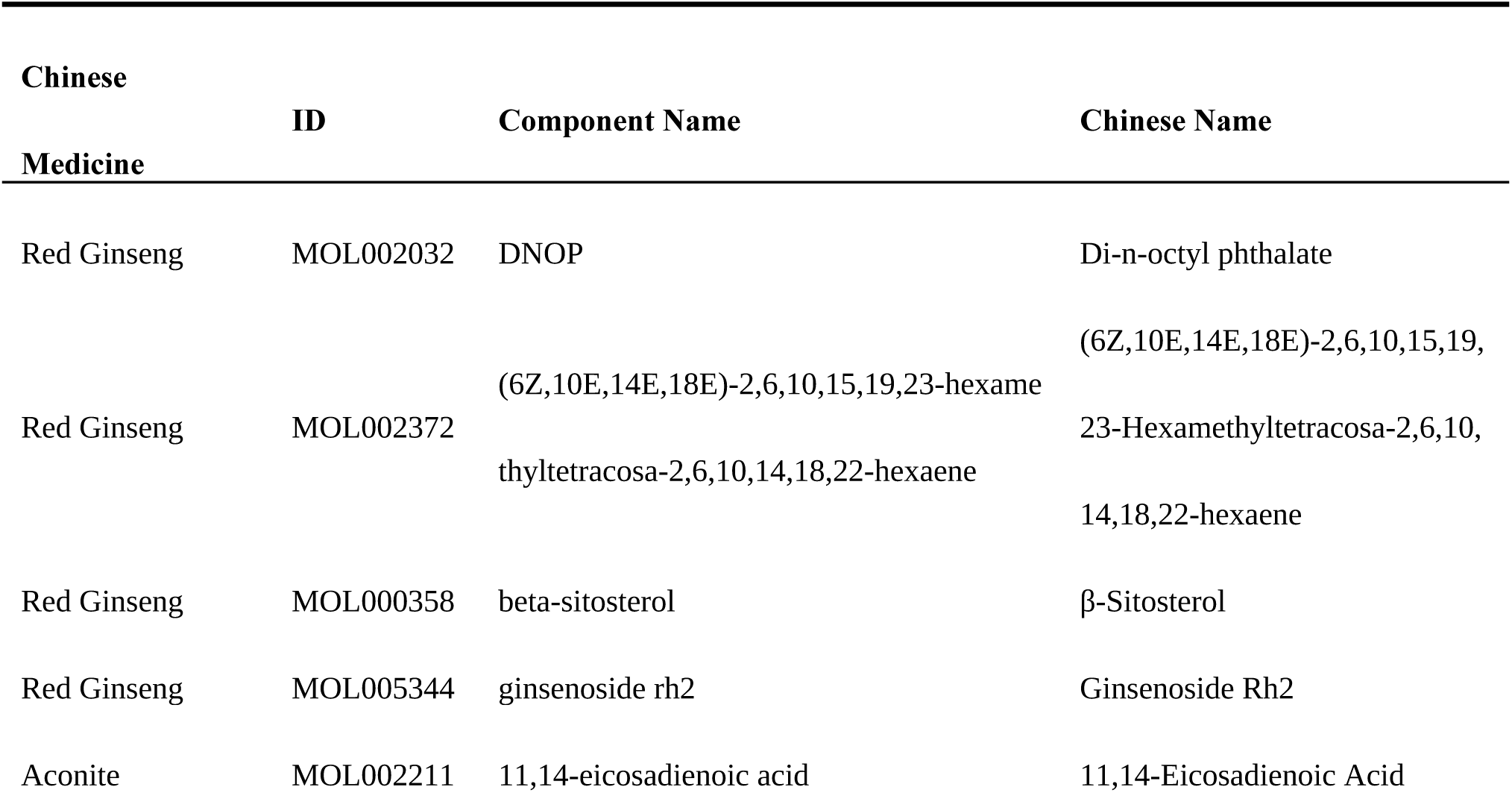
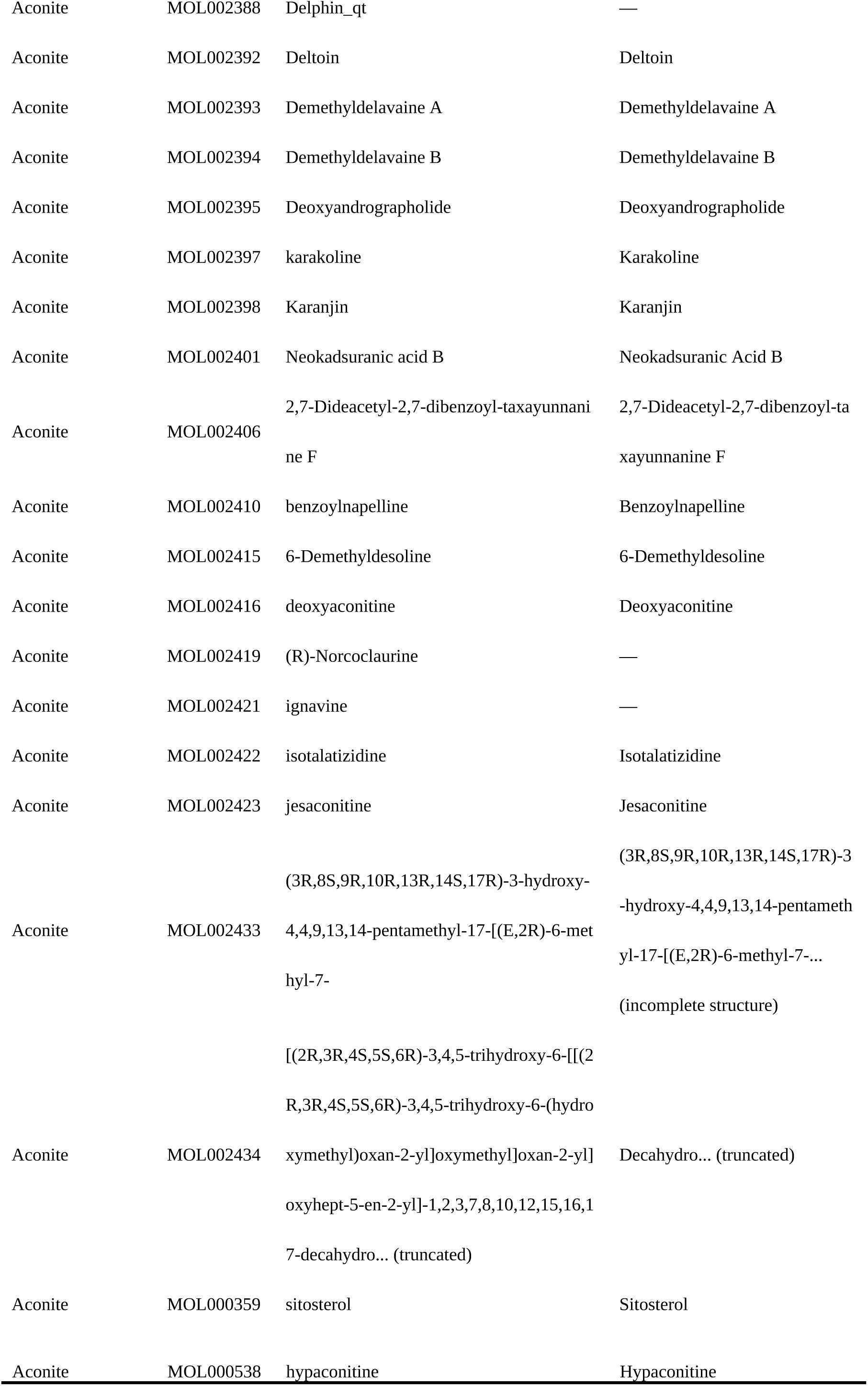
Screening of Active Components in Shenfu Injection.

### Protein-Protein Interaction (PPI) Network Analysis

The 145 common targets were imported into the STRING database to construct a PPI network. The resulting network consisted of 145 nodes and 1,962 interaction edges (Figure 2). Analysis based on betweenness centrality indicated that several targets, including Interleukin-6 (IL-6), SRC proto-oncogene (SRC), Peroxisome proliferator-activated receptor gamma (PPARG), Mitogen-activated protein kinase 3 (MAPK3), and Heat shock protein 90 alpha family class A member 1 (HSP90AA1), might be the key targets.

**Figure 2.**
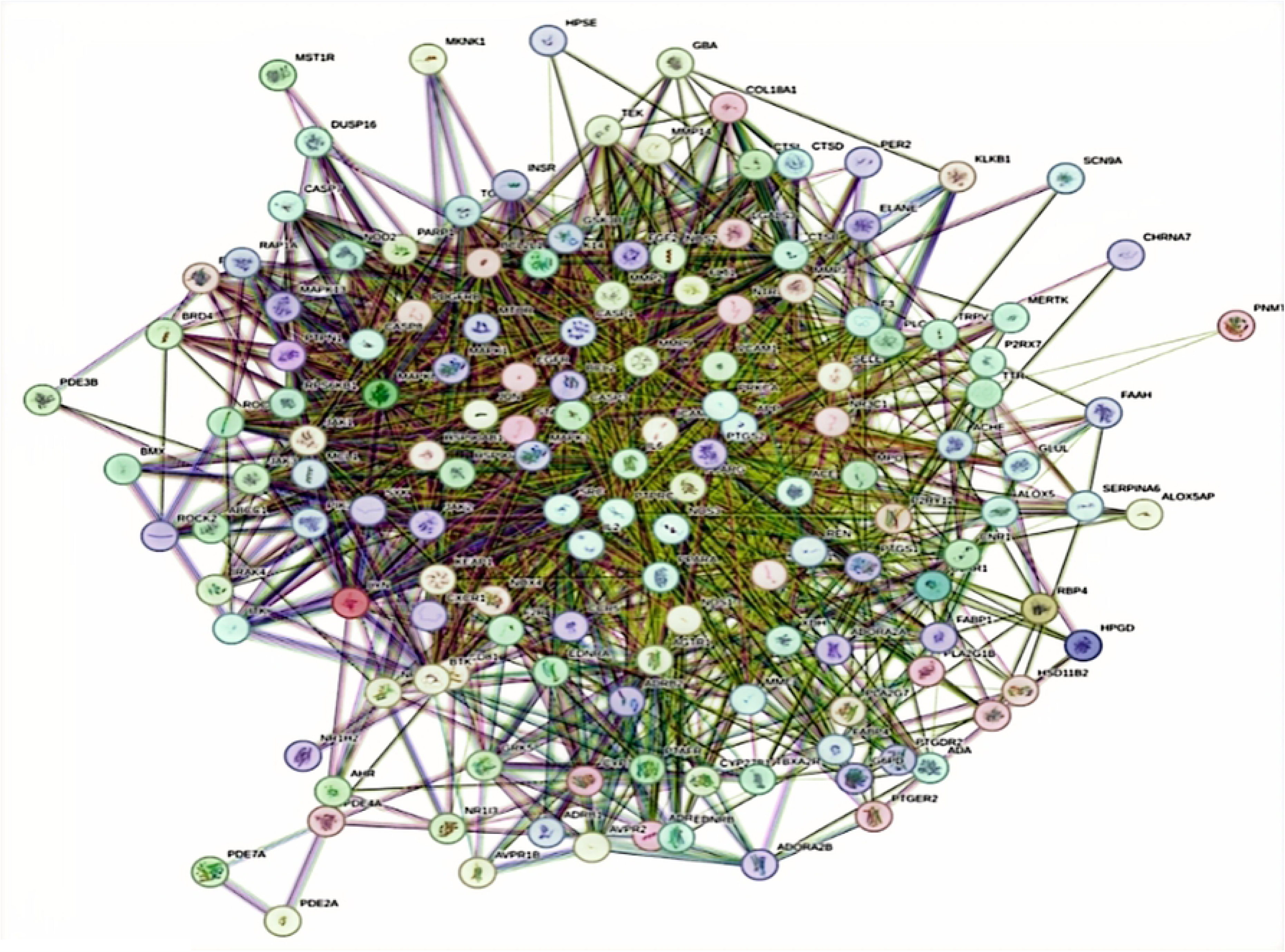
Protein-protein interaction network of common targets.

### GO and KEGG Enrichment Analysis

Gene Ontology (GO) enrichment analysis yielded 675 significant entries (P < 0.05), including 493 biological processes (BP), 69 cellular components (CC), and 113 molecular functions (MF) (Figures 3-5). The most significantly enriched BP terms were response to lipopolysaccharide, protein phosphorylation, and positive regulation of MAP kinase activity. For CC, targets were primarily localized to the plasma membrane, cell surface, integral component of plasma membrane, and membrane raft. The main MF terms included protein serine/threonine/tyrosine kinase activity, protein tyrosine kinase activity, and enzyme binding. Kyoto Encyclopedia of Genes and Genomes (KEGG) pathway analysis identified 139 significantly enriched pathways (P < 0.05) (Figure 6). The top enriched pathways included Lipid and atherosclerosis, AGE-RAGE signaling pathway in diabetic complications, Pathways in cancer, Kaposi sarcoma-associated herpesvirus infection, EGFR tyrosine kinase inhibitor resistance, TNF signaling pathway, PI3K-Akt signaling pathway, and IL-17 signaling pathway.

**Figure 3.**
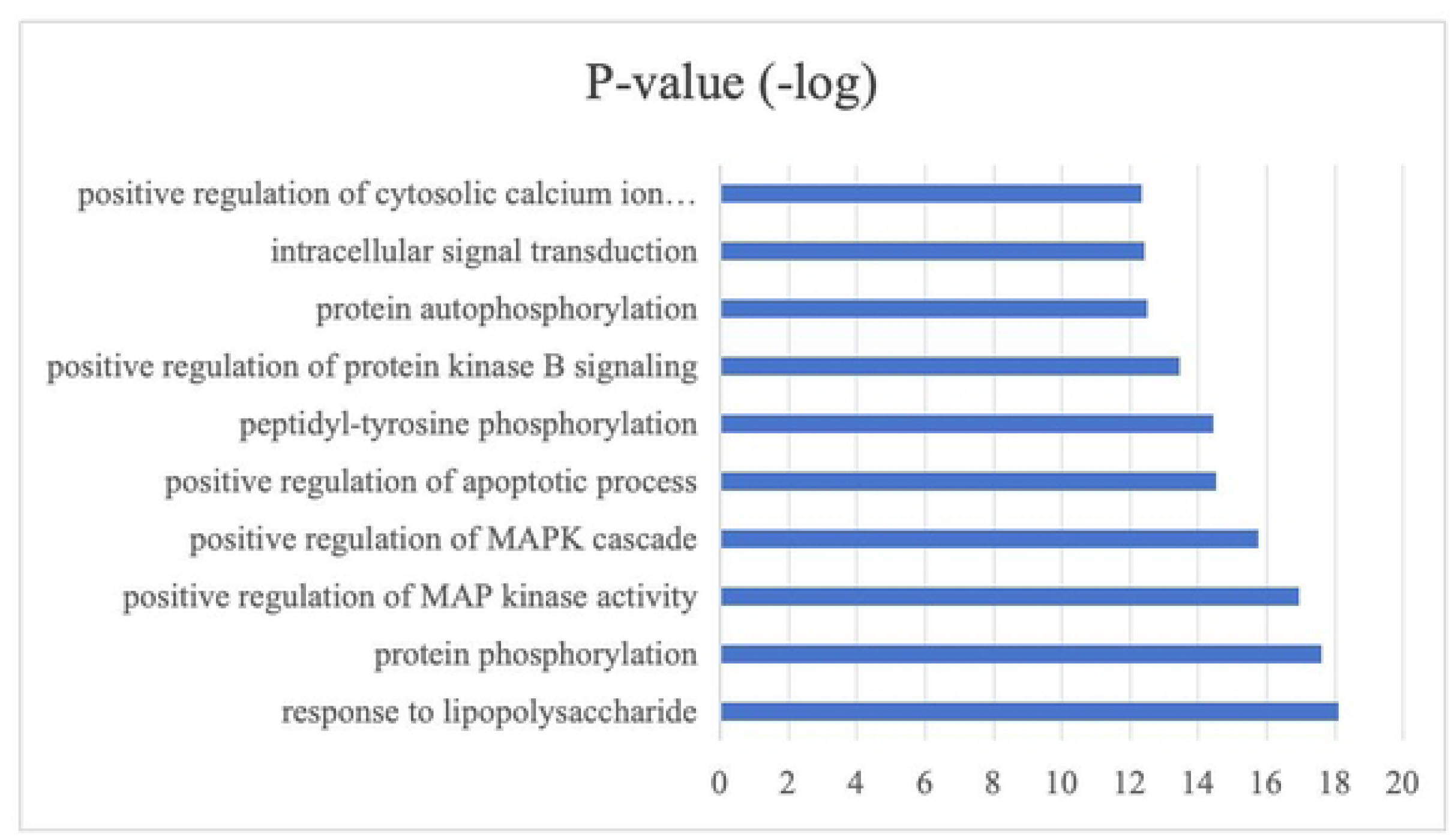
Gene Ontology enrichment analysis (Biological Process).

**Figure 4.**
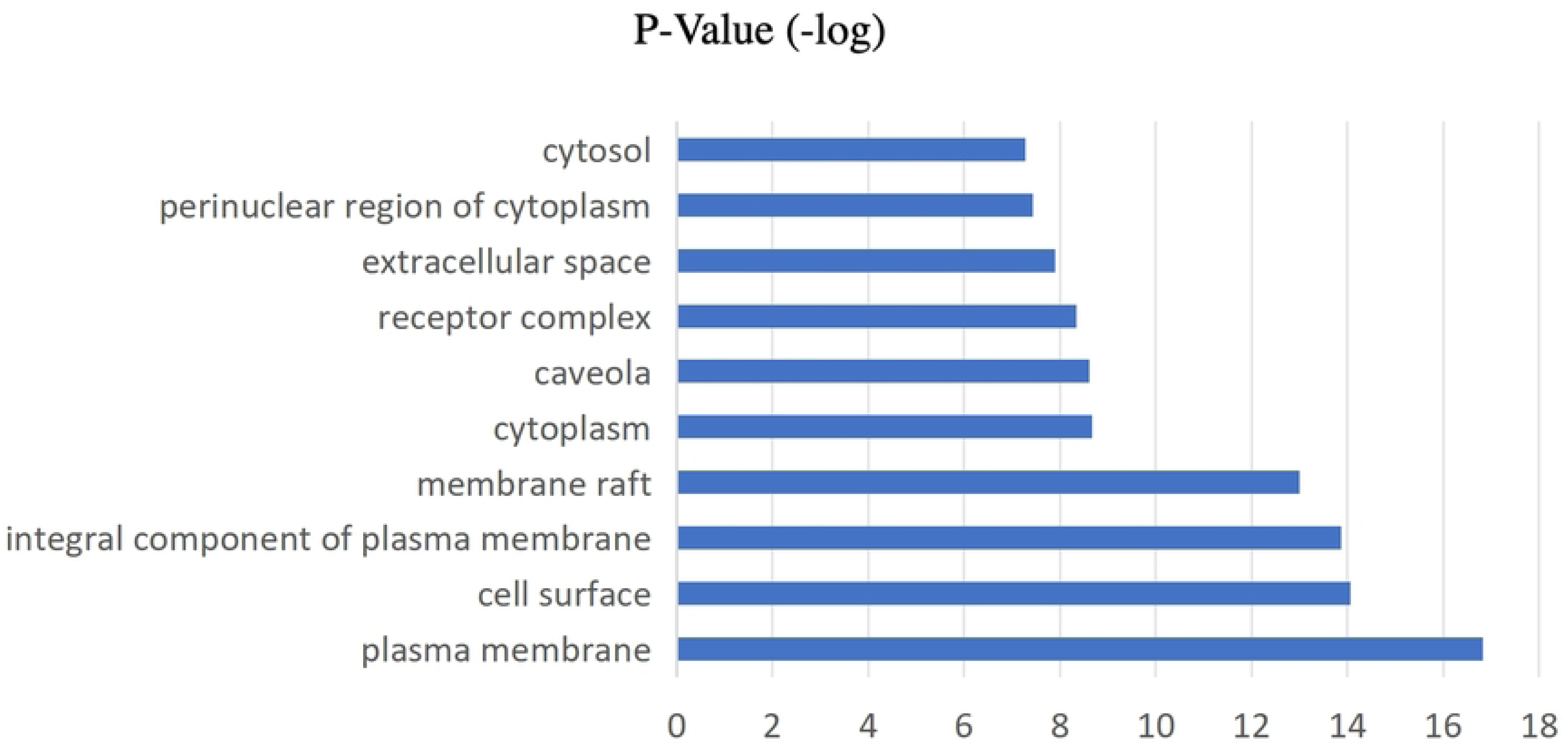
Gene Ontology enrichment analysis (Cellular Component).

**Figure 5.**
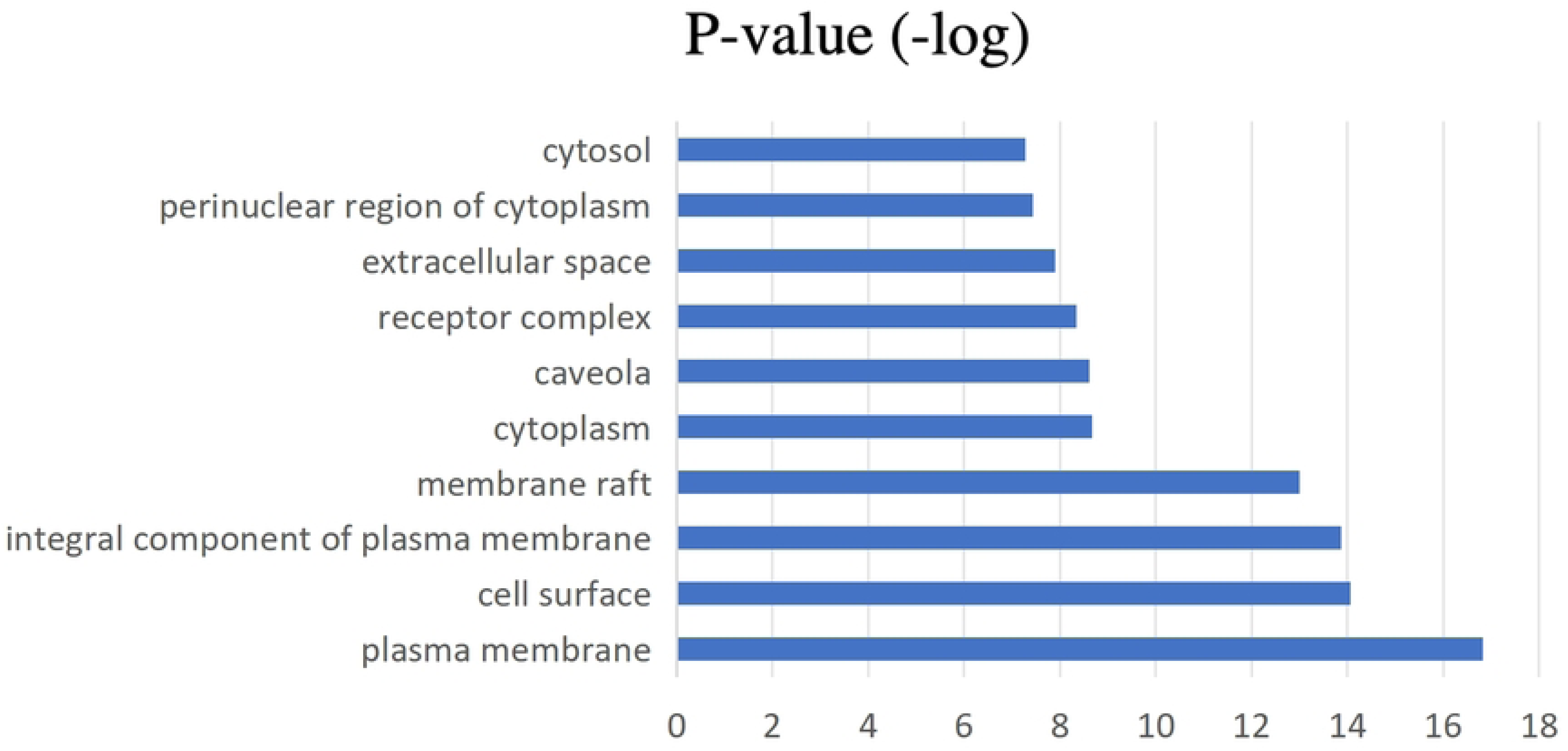
Gene Ontology enrichment analysis (Molecular Function).

**Figure 6.**
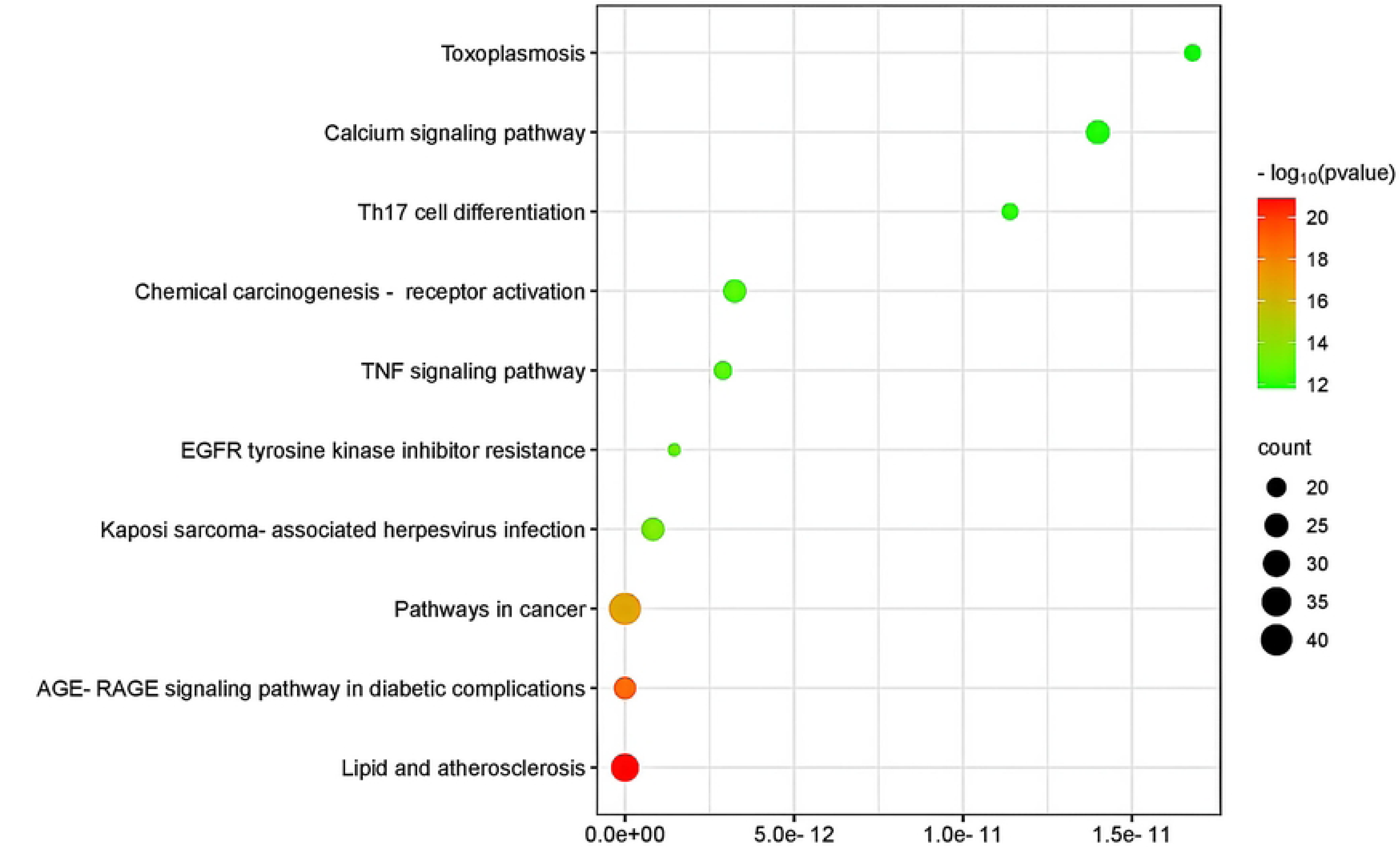
Top enriched KEGG pathways.

### Construction of the “Drug-Component-Target-Pathway” Network

The integrated network analysis (Figure 7) revealed that components with high degree values included Deltoin, Di-n-octyl phthalate (DNOP), Karanjin, Ginsenoside Rh2, Neokadsuranic acid B, and Deoxyandrographolide. Targets with high degree values were MAPK3, MAPK1, MAPK13, MAPK8, IL6, MAPK14, and MAPK10. The most connected pathways were Pathways in cancer, PI3K-Akt signaling pathway, Lipid and atherosclerosis, Chemical carcinogenesis - receptor activation, and Calcium signaling pathway.

**Figure 7.**
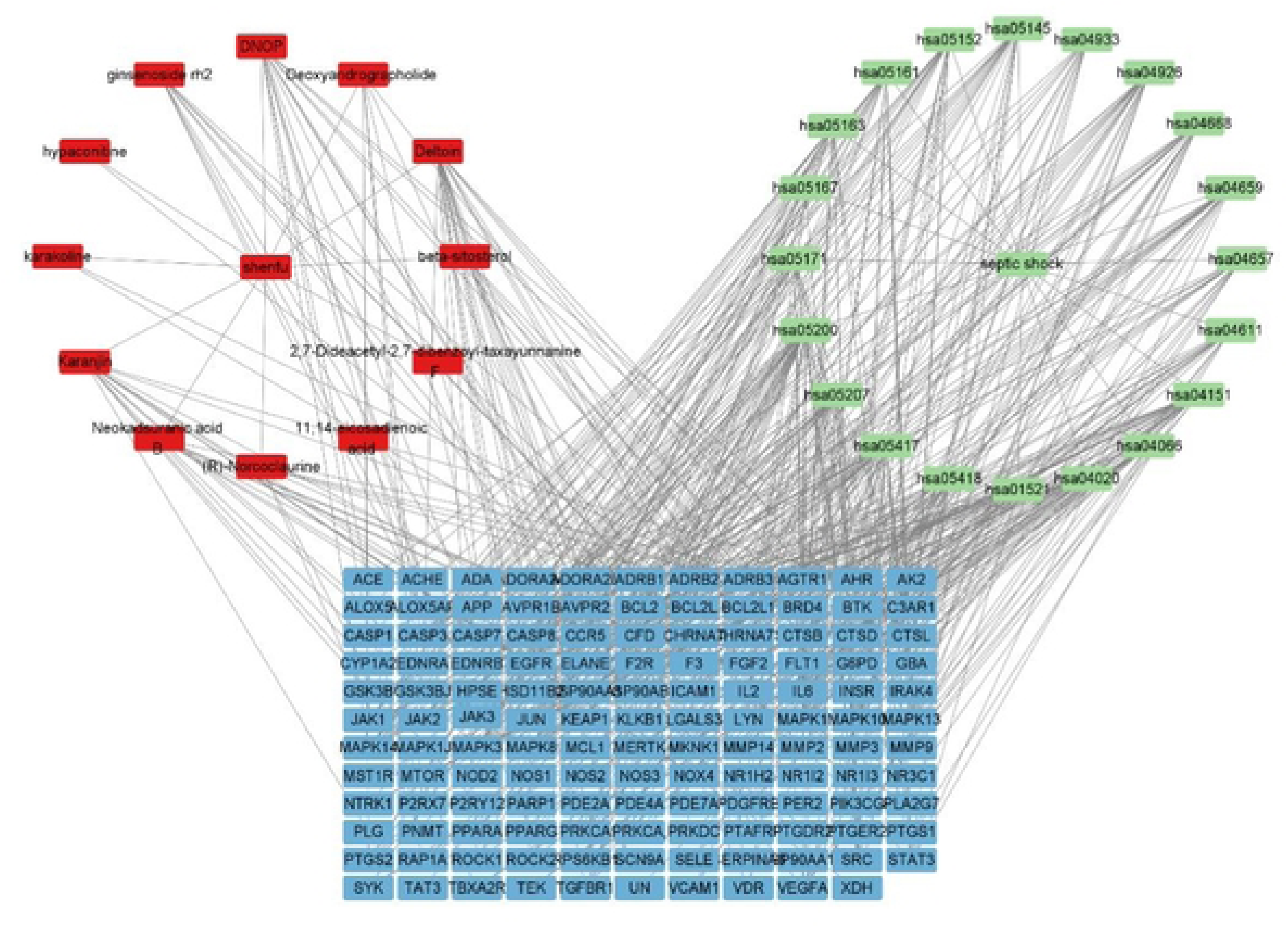
Herb-component-target-pathway network of Shenfu Injection.

### Molecular Docking Validation

Molecular docking was performed between the top 5 core target proteins (IL6, SRC, PPARG, MAPK3, HSP90AA1) and the top 5 active components (DNOP, Deltoin, Karanjin, Neokadsuranic acid B, Ginsenoside Rh2) (Figure 8). The results indicated that the key components of Shenfu Injection had good binding activity with the core targets of septic shock, as evidenced by low binding energy values.

**Figure 8.**
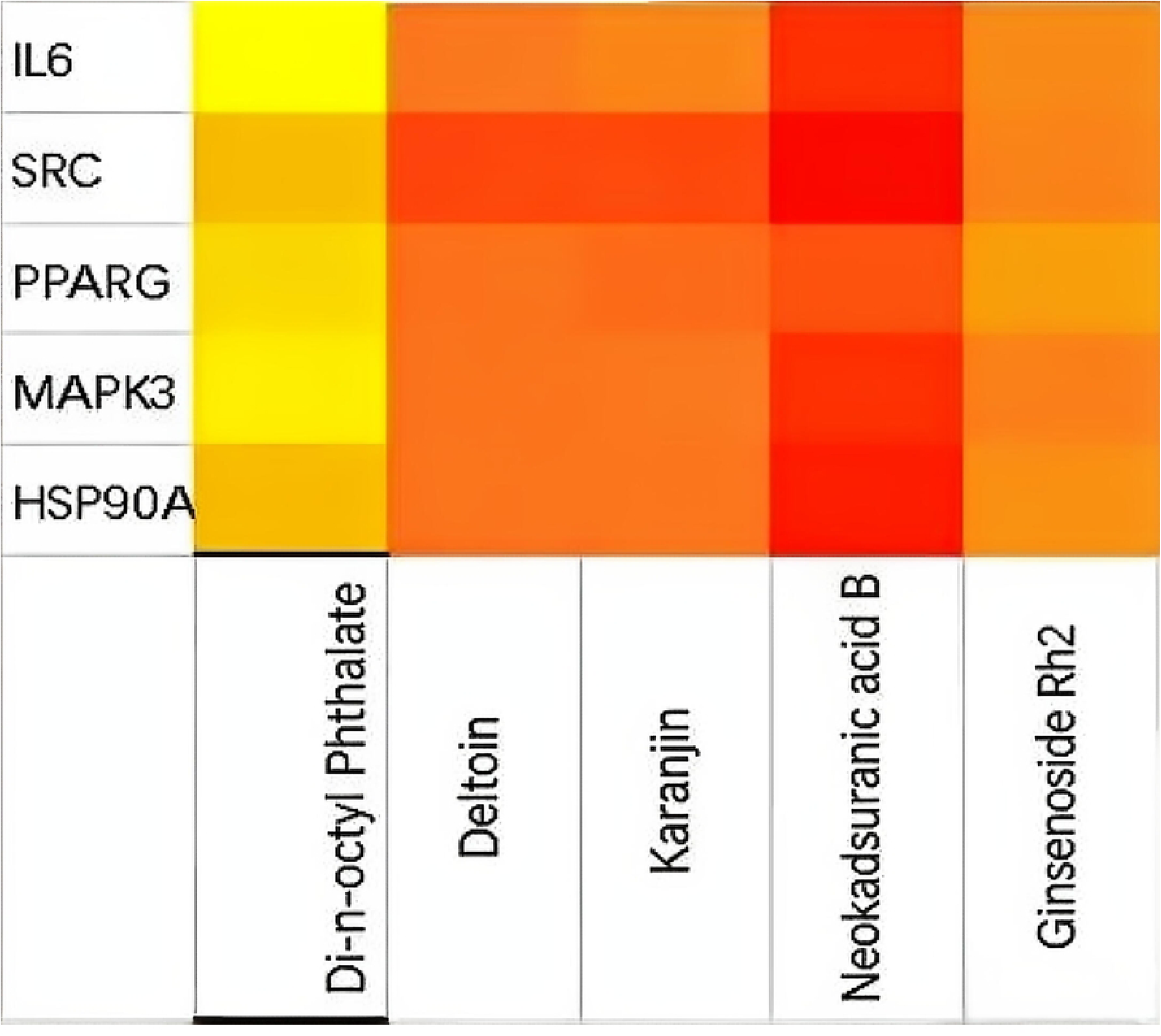
Molecular docking of core targets and active components.

## Discussion

The high mortality associated with septic shock, despite advances in standardized care, underscores the urgent need for adjunctive therapies that can modulate the dysregulated host response [16]. This integrated study, combining a prospective randomized clinical trial with a systematic network pharmacology analysis, provides compelling evidence supporting the use of Shenfu Injection (SFI) as a beneficial adjunct in septic shock management. Our findings not only confirm and extend previous clinical observations regarding SFI ’s efficacy but also, for the first time, offer a comprehensive multi-scale mechanistic framework linking its clinical benefits to underlying molecular interactions.

The primary clinical finding of our study is that adjunctive SFI therapy significantly reduced 28-day all-cause mortality from 42.5% to 20.0%. This result aligns with and strengthens the conclusions of prior meta-analyses [17] and a multi-center RCT [18], which reported mortality benefits associated with SFI. Beyond survival, our data delineate a clear pattern of accelerated clinical improvement. The SFI group demonstrated a more rapid achievement of hemodynamic stability (shorter time to target MAP, higher EGDT bundle compliance) and required a significantly shorter duration of vasopressor support. This aligns with SFI ’s known pharmacological effects of enhancing myocardial contractility and improving vascular tone [19], suggesting it may act synergistically with catecholamines to stabilize circulation more efficiently, potentially reducing the exposure to high-dose vasopressor-related complications.

More importantly, our study provides granular insights into how SFI modulates key pathophysiological axes of septic shock. The significantly greater reduction in PCT, CRP, and particularly IL-6 levels in the SFI group points towards a potent anti-inflammatory effect. IL-6 is a central mediator of the cytokine storm in sepsis, correlating with disease severity and mortality [20,21]. Its early and sustained suppression by SFI suggests a capability to temper the excessive inflammatory response, a therapeutic goal that has been elusive for many targeted anti-cytokine agents. Concurrently, the superior improvement in oxygen metabolism markers—evidenced by higher lactate clearance, higher ScvO₂, and an improved PaO₂ /FiO ₂ ratio—indicates enhanced tissue perfusion and oxygen utilization. This is consistent with previous microcirculatory studies showing SFI improves capillary perfusion [10]. The correlation between improved lactate clearance and survival is well-established, and our findings suggest SFI facilitates this critical metabolic recovery. The concomitant improvement in SOFA and APACHE II scores further corroborates the systemic benefit, translating into shorter durations of mechanical ventilation and ICU stay.

While the clinical benefits are evident, the “black box” of SFI’s mechanism has limited its broader acceptance and optimized application. Our network pharmacology analysis directly addresses this gap, moving beyond the traditional single-target paradigm to elucidate a “multi-component, multi-target, multi-pathway” mode of action, which is a hallmark of effective interventions in complex syndromes like septic shock [22,23].

The identification of 145 common targets between SFI ’s active components and septic shock pathology underscores its broad therapeutic potential. The PPI network highlighted several hub targets, including IL-6, SRC, PPARG, MAPK3 (ERK1), and HSP90AA1. The centrality of IL-6 perfectly bridges our computational prediction with clinical observation, validating it as a key mechanistic node for SFI. SRC and MAPK3 are pivotal kinases involved in intracellular signaling cascades (e.g., mediating responses to growth factors and stress), influencing inflammation, cell survival, and vascular permeability. PPARG is a nuclear receptor with potent anti-inflammatory and metabolic regulatory functions, and its modulation has been proposed as a therapeutic strategy in sepsis [24]. HSP90AA1 is a molecular chaperone critical for the stability and function of numerous client proteins, including key signaling molecules in inflammation and immune response.

KEGG pathway enrichment analysis revealed that these targets converge on several signaling pathways critically implicated in septic shock pathogenesis. The significant enrichment of the TNF signaling pathway, PI3K-Akt signaling pathway, and IL-17 signaling pathway is particularly noteworthy. These pathways are integral to the initiation and propagation of inflammatory responses, cellular apoptosis, and immune cell activation. Furthermore, the enrichment of the AGE-RAGE signaling pathway connects SFI ’s action to oxidative stress and endothelial dysfunction, common features in septic shock. The involvement of the Lipid and atherosclerosis pathway, while seemingly unrelated, may reflect shared mechanisms of inflammation and endothelial injury. The Pathways in cancer enrichment, a common finding in network pharmacology, likely signifies SFI’s broad impact on core cellular processes such as proliferation, apoptosis, and survival, which are also disrupted in septic organs.

The constructed “Drug-Component-Target-Pathway” network visually encapsulates SFI’s holistic strategy. Key components like Ginsenoside Rh2 (from ginseng) and Deltoin (from aconite) were connected to multiple core targets and pathways.

Molecular docking provided preliminary validation, showing strong binding affinities between these components and hub targets like IL-6 and MAPK3. This suggests that the clinical anti-inflammatory effect may be directly mediated by compounds like Ginsenoside Rh2 interfering with IL-6 signaling, while the improvement in cellular metabolism and survival could be linked to the modulation of the PI3K-Akt pathway via other components.

The principal innovation of this study lies in the concurrent and integrative application of clinical research and network pharmacology. This approach allows us to propose a coherent narrative: SFI, through its diverse bioactive components (e.g., ginsenosides, alkaloids), simultaneously engages multiple key targets (e.g., IL-6, MAPK3, PPARG). This concerted action dampens overactive inflammatory pathways (TNF, IL-17), promotes cellular survival signals (PI3K-Akt), and mitigates endothelial and metabolic dysfunction, as reflected in the rapid downregulation of systemic inflammation (lower IL-6, PCT) and the improvement in tissue oxygen metabolism (higher lactate clearance) observed clinically. This network-based effect may explain why SFI demonstrates efficacy where single-target anti-inflammatory biologics have largely failed in sepsis trials—it restores balance across the interconnected biological network rather than blocking a single point [25].

Our study has several limitations that warrant acknowledgment. First, the clinical trial was a single-center, open-label study, which may introduce potential bias. Although baseline characteristics were balanced, a larger, multi-center, double-blind, placebo-controlled trial is needed to conclusively confirm our findings. Second, the network pharmacology predictions, though validated by molecular docking, are primarily computational. Experimental validation using in vitro (e.g., LPS-stimulated macrophages) and in vivo (e.g., septic animal models) studies is essential to confirm the modulation of the predicted core targets (e.g., phosphorylation status of MAPK3, activity of SRC) and pathways. Third, the pharmacokinetics and tissue distribution of the identified active components in critically ill patients remain unclear and should be investigated. Future research should focus on: 1) Conducting rigorous multicenter RCTs with biomarker-stratified patient subgroups to identify who benefits most from SFI; 2) Employing systems biology approaches like transcriptomics and proteomics on patient blood samples pre- and post-SFI treatment to empirically map the in vivo network response; 3) Isolating and studying the synergistic interactions between the top-predicted component pairs; 4) Exploring the potential of SFI’s core components as leads for designing novel multi-target therapeutic agents.

## Conclusion

In conclusion, this integrative study demonstrates that adjunctive therapy with Shenfu Injection in patients with septic shock is associated with a significant reduction in 28-day mortality, accelerated resolution of inflammation and tissue hypoxia, and improved clinical recovery. The network pharmacology analysis provides a plausible and comprehensive molecular blueprint for these effects, characterizing SFI as a multi-target agent capable of modulating a network of key targets and pathways central to septic shock pathogenesis, including inflammation, cellular signaling, and metabolic stress. These findings strengthen the evidence base for SFI as a valuable complementary therapy in septic shock and exemplify a powerful paradigm for elucidating the mechanism of action of complex natural products in critical illness.

## Data Availability

All relevant data underlying the results presented in the study are contained within the manuscript and its Supporting Information files. For further inquiries regarding the original data, researchers can contact the corresponding author (Yahui Shi).

## Acknowledgments

Not applicable.

## Author Contributions

Conception and design of the research: B.Z., Y.S. Acquisition of data: B.Z., Y.S., X.Z. Analysis and interpretation of the data: Y.T., Q.L. Statistical analysis: Y.T. Obtaining financing: Y.S. Writing of the manuscript: B.Z. Critical revision of the manuscript for intellectual content: Y.S., Y.T., Q.L., X.Z.

## Ethics Statement

The studies involving human participants were reviewed and approved by the Ethics Committee of Cangzhou Hospital of Integrated Traditional Chinese and Western Medicine-Hebei (Approval No. CZX2023-KY-027.1). Written informed consent was obtained from all participants or their legal surrogates.

## Data availability statement

The original contributions presented in the study are included in the article/supplementary material, further inquiries can be directed to the corresponding author/s.

## Abbreviations List

SFI: Shenfu Injection
TCM: Traditional Chinese Medicine
MODS: Multiple Organ Dysfunction Syndrome
MAP: Mean Arterial Pressure
SOFA: Sequential Organ Failure Assessment
APACHE II: Acute Physiology and Chronic Health Evaluation II
PCT: Procalcitonin
CRP: C-reactive Protein
IL-6: Interleukin-6
EGDT: Early Goal-Directed Therapy
ScvO₂: Central Venous Oxygen Saturation
ICU: Intensive Care Unit
EICU: Emergency Intensive Care Unit
RCT: Randomized Controlled Trial
OB: Oral Bioavailability
DL: Drug-Likeness
PPI: Protein-Protein Interaction
GO: Gene Ontology
KEGG: Kyoto Encyclopedia of Genes and Genomes
BP: Biological Process
CC: Cellular Component
MF: Molecular Function
TNF: Tumor Necrosis Factor
PI3K: Phosphatidylinositol 3-Kinase
Akt: Protein Kinase B
MAPK3: Mitogen-Activated Protein Kinase 3
SRC: Proto-oncogene Tyrosine-Protein Kinase Src
PPARG: Peroxisome Proliferator-Activated Receptor Gamma
HSP90AA1: Heat Shock Protein 90 Alpha Family Class A Member 1
OR: Odds Ratio
CI: Confidence Interval
SD: Standard Deviation
IQR: Interquartile Range
PDB: Protein Data Bank

## Reference

[1] Meyer NJ, Prescott HC. Sepsis and Septic Shock. N Engl J Med, 2024, 391(22):2133–2146. DOI:10.1056/NEJMra2403213.

[2] Meyhoff TS, Hjortrup PB, Wetterslev J, et al. Restriction of Intravenous Fluid in ICU Patients with Septic Shock. N Engl J Med, 2022, 386(26):2459–2470. DOI:10.1056/NEJMoa2202707.

[3] Delaney A, Borges-Sa M, Chew MS, et al. Current standard of care for septic shock. Intensive Care Med, 2025. DOI:10.1007/s00134-025-08211-6.

[4] Bakker J, Kattan E, Annane D, et al. Current practice and evolving concepts in septic shock resuscitation. Intensive Care Med, 2022, 48(2):148–163. DOI:10.1007/s00134-021-06595-9.

[5] Pfortmueller CA, Dabrowski W, Wise R, et al. Fluid accumulation syndrome in sepsis and septic shock: pathophysiology, relevance and treatment-a comprehensive review. Ann Intensive Care, 2024, 14(1):115. DOI:10.1186/s13613-024-01336-9.

[6] Cao M, Shi M, Zhou B, Jiang H. An overview of the mechanisms and potential roles of extracellular vesicles in septic shock. Front Immunol, 2024, 14:1324253. DOI:10.3389/fimmu.2023.1324253.

[7] Yu Y, Zhu C, Hong Y, et al. Effectiveness of anisodamine for the treatment of critically ill patients with septic shock: a multicentre randomized controlled trial. Crit Care, 2021, 25(1):349. DOI:10.1186/s13054-021-03774-4.

[8] Wang M, Liu H, Chen Y, et al. Guideline on treating community-acquired pneumonia with Chinese patent medicines. Pharmacol Res, 2023, 196:106919. DOI:10.1016/j.phrs.2023.106919.

[9] Tian R, Li R, Chen Y, et al. Shenfu injection ameliorates endotoxemia-associated endothelial dysfunction and organ injury via inhibiting PI3K/Akt-mediated glycolysis. J Ethnopharmacol, 2024, 335:118634. DOI:10.1016/j.jep.2024.118634.

[10] Zhang Z, Chen L, Sun B, et al. Identifying septic shock subgroups to tailor fluid strategies through multi-omics integration. Nat Commun, 2024, 15:9028. Doi: 10.1038/s41467-024-53239-9.

[11] Wang X, Guo R, Guo Y, et al. Rationale and design of the RESTORE trial: A multicenter, randomized, double-blinded, parallel-group, placebo-controlled trial to evaluate the effect of Shenfu injection on myocardial injury in STEMI patients after primary PCI. Am Heart J, 2023, 260:9–17. DOI:10.1016/j.ahj.2023.02.005.

[12] Li X, Huang F, Zhu L, et al. Effects of combination therapy with Shenfu Injection in critically ill patients with septic shock receiving mechanical ventilation: A multicentric, real-world study. Front Pharmacol, 2022, 13:1041326. DOI:10.3389/fphar.2022.1041326.

[13] Yang B, Wang S, Yang Y, Wang Y. Toxicity and safety profile evaluation of Shenfu injection in a murine sepsis model. J Ethnopharmacol, 2025, 337(Pt 2):118903. DOI:10.1016/j.jep.2024.118903.

[14] Niu L, Xiao L, Zhang X, et al. Comparative Efficacy of Chinese Herbal Injections for Treating Severe Pneumonia: A Systematic Review and Bayesian Network Meta-Analysis of Randomized Controlled Trials. Front Pharmacol, 2022, 12:743486. DOI:10.3389/fphar.2021.743486.

[15] Huang P, Guo Y, Hu X, et al. Mechanism of Shenfu injection in suppressing inflammation and preventing sepsis-induced apoptosis in murine cardiomyocytes based on network pharmacology and experimental validation. J Ethnopharmacol, 2024, 322:117599. DOI:10.1016/j.jep.2023.117599.

[16] Seymour CW, Liu VX, Iwashyna TJ, et al. Assessment of Clinical Criteria for Sepsis: For the Third International Consensus Definitions for Sepsis and Septic Shock (Sepsis-3). JAMA, 2016, 315(8):762–774. DOI:10.1001/jama.2016.0288.

[17] Liao J, Qin C, Wang Z, et al. Effect of shenfu injection in patients with septic shock: A systemic review and meta-analysis for randomized clinical trials. J Ethnopharmacol, 2024, 320:117431. DOI:10.1016/j.jep.2023.117431.

[18] Li Y, Zhang X, Lin P, et al. Effects of Shenfu Injection in the Treatment of Septic Shock Patients: A Multicenter, Controlled, Randomized, Open-Label Trial. Evid Based Complement Alternat Med, 2016, 2016:2565169. DOI:10.1155/2016/2565169.

[19] Huang P, Chen Y, Zhang H, et al. Comparative Efficacy of Chinese Herbal Injections for Septic Shock: A Bayesian Network Meta-Analysis of Randomized Controlled Trials. Front Pharmacol, 2022, 13:850221. DOI:10.3389/fphar.2022.850221.

[20] Chen J, Ding W, Zhang Z, et al. Shenfu injection targets the PI3K-AKT pathway to regulate autophagy and apoptosis in acute respiratory distress syndrome caused by sepsis. Phytomedicine, 2024, 129:155627. DOI:10.1016/j.phymed.2024.155627.

[21] Ji Y, Song H, Li L. Traditional Chinese medicine for sepsis: advancing from evidence to innovative drug discovery. Crit Care, 2025, 29(1):193. DOI:10.1186/s13054-025-05441-4.

[22] Yang R, Hu C, Zhuo Y, et al. Comparative efficacy of Chinese tonic medicines for treating sepsis or septic shock: A systematic review and Bayesian network meta-analysis of randomized controlled trials. Phytomedicine, 2025, 136:156295. DOI:10.1016/j.phymed.2024.156295.

[23] Liu X, Ding H, Chen M, et al. Shenfu Injection Mediated NLRP3/Caspase 1 Through (R)-Norcoclaurinee Alleviates Sepsis-Induced Cognitive Dysfunction. J Inflamm Res, 2024, 17:7295–7310. DOI:10.2147/JIR.S481171.

[24] Zhou Y, Zhu Y, Wu Y, et al. 4-phenylbutyric acid improves sepsis-induced cardiac dysfunction by modulating amino acid metabolism and lipid metabolism via Comt/Ptgs2/Ppara. Metabolomics, 2024, 20(3):46. DOI:10.1007/s11306-024-02112-3.

[25] Xiao L, Niu L, Xu X, et al. Comparative Efficacy of Tonic Chinese Herbal Injections for Treating Sepsis or Septic Shock: A Systematic Review and Bayesian Network Meta-Analysis of Randomized Controlled Trials. Front Pharmacol, 2022, 13:830030. DOI:10.3389/fphar.2022.830030.

